# Age-targeted dose allocation can halve COVID-19 vaccine requirements

**DOI:** 10.1101/2020.10.08.20208108

**Authors:** Michael T. Meehan, Daniel G. Cocks, Jamie M. Caldwell, James M. Trauer, Adeshina I. Adekunle, Romain R. Ragonnet, Emma S. McBryde

**Affiliations:** Australian Institute of Tropical Health and Medicine, James Cook University, Townsville, 4811, Australia; Research School of Physics, Australian National University, Canberra, Australia; Department of Biology, University of Hawaii at Manoa, Hawaii, United States of America; Epidemiological Modelling Unit, School of Public Health and Preventive Medicine, Monash University, Melbourne, Australia

## Abstract

In anticipation of COVID-19 vaccine deployment, we use an age-structured mathematical model to investigate the benefits of optimizing age-specific dose allocation to suppress the transmission, morbidity and mortality of SARS-CoV-2 and the associated disease, COVID-19. To minimize transmission, we find that the highest priority individuals across 179 countries are typically those between 30 and 59 years of age because of their high contact rates and higher risk of infection and disease. Conversely, morbidity and mortality are initially most effectively reduced by targeting 60+ year olds who are more likely to experience severe disease. However, when population-level coverage is sufficient — such that herd immunity can be achieved through targeted dose allocation — prioritizing middle-aged individuals becomes the most effective strategy to minimize hospitalizations and deaths. For each metric considered, we show that optimizing the allocation of vaccine doses can more than double their effectiveness.

## Main

Several promising vaccines for severe acute respiratory syndrome coronavirus-2 (SARS-CoV-2), the infection that causes coronavirus disease 19 (COVID-19), are now awaiting approval^1,2^ with early shipments of doses anticipated in 2021. Although the leading candidates have shown remarkable effectiveness (>90%) in protecting vaccine recipients from contracting the disease, limited global supplies necessitate that available doses are allocated as efficiently as possible: targeting either elderly individuals at greatest risk of infection and disease, or middle-aged individuals most responsible for ongoing transmission.

At present, the choice to target transmitters or focus on the vulnerable likely depends on the degree of control attainable by each host country. Whilst SARS-CoV-2 has a relatively low case-fatality rate (compared with e.g., SARS-CoV-1, Middle East Respiratory Syndrome)^3^, it is highly transmissible (e.g., basic reproduction number estimated at *R*_0_ = 2 − 3)^4^ and notoriously difficult to contain. In contrast with SARS-CoV-1, transmission of SARS-CoV-2 is partly driven by asymptomatic and presymptomatic individuals^5^, with the former accounting for up to 30% of all infections by some estimates^6^. Such “silent transmission” severely complicates infection control as it limits both (i) the capacity of non-pharmaceutical interventions such as case isolation and contact tracing to identify carriers and contain the spread of infection; and (ii) the potential impact on transmission of vaccines that prevent symptomatic disease only, since asymptomatic individuals can remain highly infectious. Ideally, a vaccine with sufficient efficacy and coverage would allow communities to achieve herd immunity — where the immune fraction of the population yields an effective reproduction number, *R*_eff_ *<* 1; however, limited doses (with sub-optimal efficacy) may force many vaccination programs to forgo elimination as a realistic target and prioritize reductions in COVID-19 morbidity and mortality.

In the absence of pre-existing immunity, the minimum population-level vaccination coverage required to achieve herd immunity (under the assumption of homogeneous mixing and susceptibility across the population) is (1 − 1*/R*_0_)*/e*; where the vaccine efficacy, *e*, is included because even the most effective vaccines only confer partial protection — particularly amongst the elderly. Given an *R*_0_ of 2 – 3, this formula predicts SARS-CoV-2 herd immunity thresholds at between 50 and 67%, which equates to approximately 56 - 75% population-wide vaccination coverage for a vaccine that is 90% effective. However, recent studies incorporating population heterogeneity have suggested that SARS-CoV-2 herd immunity thresholds may be considerably lower than naïve estimates^8−11^. This is due to the fact that young-to-middle-age groups have high contact rates combined with high infectiousness and susceptibility to infection. Removing these persons from the susceptible pool would therefore have disproportionate effects on the transmission potential. These observations are crucial for vaccination strategies, as they imply that the age-specific characteristics of both social mixing and SARS-CoV-2 infection could be leveraged to increase the efficiency of vaccine campaigns — potentially restoring vaccine-induced herd immunity as a realistic goal.

In this study we investigate the contrasting objectives of minimizing SARS-CoV-2 transmission and reducing COVID-19 morbidity and mortality by using an age-stratified SARS-CoV-2 transmission model. The model, which incorporates age-dependent susceptibility and disease severity calibrated to COVID-19 patient data^12^, is used to estimate the age-specific transmission rates among infected individuals, and the cumulative number of hospitalized cases and deaths in 179 countries. We then use the efficacy profiles of several leading vaccine candidates (alongside those proposed by the World Health Organization^13^) to identify the optimal age-specific allocation of vaccine doses to limit SARS-CoV-2 transmission and COVID-19-related morbidity and mortality by: 1) minimizing the effective reproduction number, *R*_eff_, for a fixed number of available vaccine doses (categorized according to the fraction of the population that could receive a full vaccination course, i.e., the population-level coverage); 2) minimizing the number of individuals that require hospitalization as a result of infection during the course of the epidemic; 3) minimizing the number of deaths; and 4) minimizing the number of doses required to suppress transmission and achieve elimination (*R*_eff_ *<* 1).

We consider two separate modes of action for vaccine candidates: those that reduce the recipient’s susceptibility to infection; and those that protect against symptomatic disease in persons who become infected. In each case, vaccine doses are allocated independently among individuals divided into 10-year age bands from 0 − 9 up to 70+ years of age.

At baseline, we assume that the vaccine is 90% effective^13^ across all age groups and that both individual susceptibility and risk of symptomatic disease are age-dependent (see Methods). Variations to these assumptions are investigated through sensitivity analysis (Supplement).

To demonstrate the breadth of optimal solutions that can be obtained across different regions, we provide detailed results for India, China and the United Kingdom (UK). These countries span low- to high-income settings and exhibit diverse population demographic structures, contact networks — where the latter two (China^14^ and the UK^15^) have pre-COVID-19 contact survey data available — and seroprevalences. We also provide the global distribution of minimum vaccination coverage required to achieve herd immunity. Detailed, age-specific breakdowns for the remaining 176 countries appear in the Supplement.

Moreover, since the extent of infection-induced immunity generated by epidemic waves prior to the introduction of vaccination is likely to be comparable to the estimated herd immunity threshold in some settings (e.g., the UK)^17^, we extend our analysis by exploring how the presence of substantial pre-existing immunity modifies the optimal amount and distribution of doses required.

## Results

### Age-specific transmission rates

The different age distributions (Fig. 1**A**-**C**) and contact patterns in different countries drive highly heterogeneous transmission rates (Fig. 1**D**-**F**) among age groups and settings. Transmission in low- (e.g., India) and middle-income countries, which typically have bottom-heavy age pyramids, is predominately driven by younger age groups (0-29 years of age), particularly between individuals of the same age and from younger to older ages. However, there is often some additional inter-generational transmission generated by those between 20 and 59 years of age (Fig. 1**A**,**D**). Conversely, in China where high numbers of daily contacts were recorded among those aged 60 and above^14^, transmission is much more intensely concentrated in the older age groups. In high-income settings (e.g., the UK), we see that in addition to the intense within-age-group transmission among those aged 19 years and below (which is similar to low-income settings), there is considerably more inter-generational transmission generated by those individuals aged between 20 and 69 years, which becomes even more intense for regions with top-heavy age distributions such as Japan and Hong Kong.

**Figure 1.**
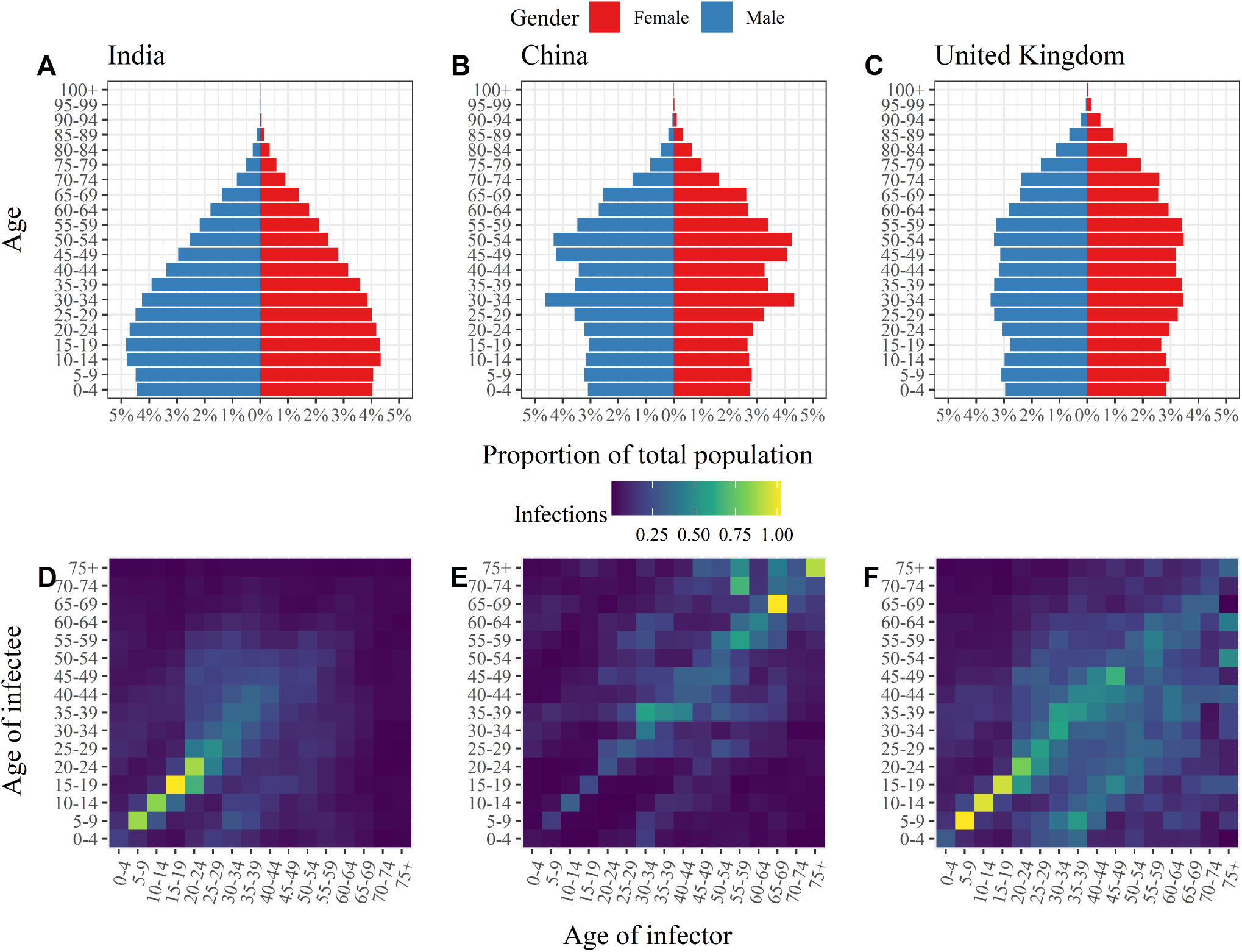
Age distributions and age-specific transmission rates. Population pyramids (**Panels A-C**) and transmission matrices (**Panels D-F**) for India (left), China (middle) and the United Kingdom (right). In **Panels D**-**F** the colouring of the *i*th row and *j*th column of the transmission matrices represents the average number of infections in age group *i* generated by an individual in age group *j* over the course of an infectious episode with COVID-19. The elements of each matrix have been rescaled such that the maximal eigenvalues of the respective transmission matrices match the basic reproduction numbers (*R*_0_) estimated by Abbott et al.^16^, namely India: 2.2; China: 2.6; and the United Kingdom: 2.3.

### Minimizing transmission, hospitalizations and deaths with a fixed number of doses

Targeted vaccination dramatically reduces the transmission rate (*R*_eff_) and cumulative number of hospitalizations and deaths relative to vaccination programs that uniformly distribute doses across the population (Fig. 2). Differences between the uniform and targeted strategies are greatest at intermediate levels of population-level coverage (e.g., 40-60%) as results under the two strategies converge when coverage approaches 0 or 100%.

**Figure 2.**
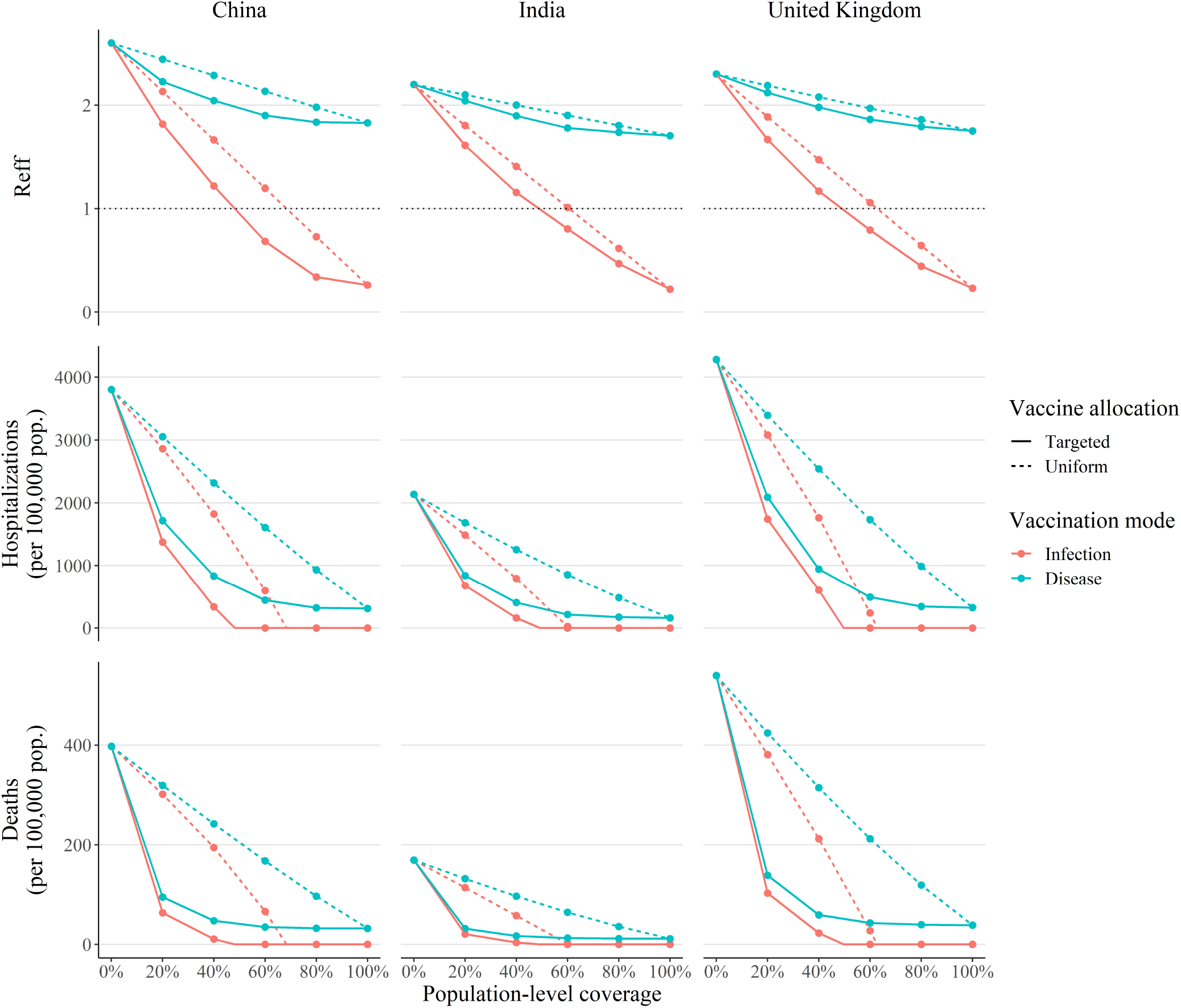
Reduction in transmission, hospitalizations and deaths under optimized vaccine allocation. Effective reproduction number *R*_eff_ (top), cumulative hospitalizations (middle) and deaths (bottom), as a function of the population-level vaccination coverage when allocation is targeted towards priority age groups (solid lines) and the vaccine protects against infection (red) and disease (blue) for India (left), China (middle) and the United Kingdom (right). For comparison, we also plot the analogous reduction in the three optimization targets when vaccine doses are allocated uniformly across age groups (dashed lines). In the top panel row, the herd immunity threshold (*R*_eff_<1) is indicated by the horizontal dotted black line.

Countries with contacts concentrated among a limited number of age groups (e.g., China, Fig. 2**B**) exhibit greater reductions in each of the three optimization targets considered when doses are allocated to priority age groups, as do countries with higher median ages (e.g., the UK, Fig. 2**C**) because older populations with rectangular age distributions possess greater scope for targeted dose allocation.

Vaccines that reduce susceptibility to infection more effectively suppress transmission, hospitalizations and deaths than those that prevent symptomatic disease among infected individuals (Fig. 2). The reduction in impact is greatest for transmission, where vaccines that prevent severe disease fail to reach the herd immunity threshold in almost all settings considered (the exceptions being countries with baseline *R*_0_ values very close to one, e.g., Samoa). Nevertheless, the ability to achieve herd immunity enables infection-preventing vaccines to reduce hospitalizations and deaths to zero (the x-intercepts in the middle and bottom rows of Fig. 2).

Looking at the age-specific dose allocation under optimized vaccination (Fig. 3), for infection-preventing vaccines (**Panel A**) we observe that whilst vaccinating individuals between 30 and 49 years of age is the most efficient way to minimize transmission (top row); when doses are limited (e.g., 20% population-level coverage), targeting older individuals (60+ years of age) is the most efficient way to minimize hospitalizations (middle row) and deaths (bottom row). However, as the population-level coverage increases towards the optimized herd-immunity threshold (where elimination is feasible) middle-aged individuals rapidly succeed the elderly as the highest priority age groups. Notable exceptions to this pattern include China (Fig. 3, left column) where the high number of contacts among individuals over the age of 60 ensure the elderly (i.e., 60+ year olds) remain the first group vaccinated regardless of the optimization objective or population coverage. Alternatively, for disease-preventing vaccines (which typically fail ever to achieve herd immunity) older age groups consistently receive top priority for hospitalizations and deaths, whereas the results for transmission are equivalent to those for infection-preventing vaccines.

**Figure 3.**
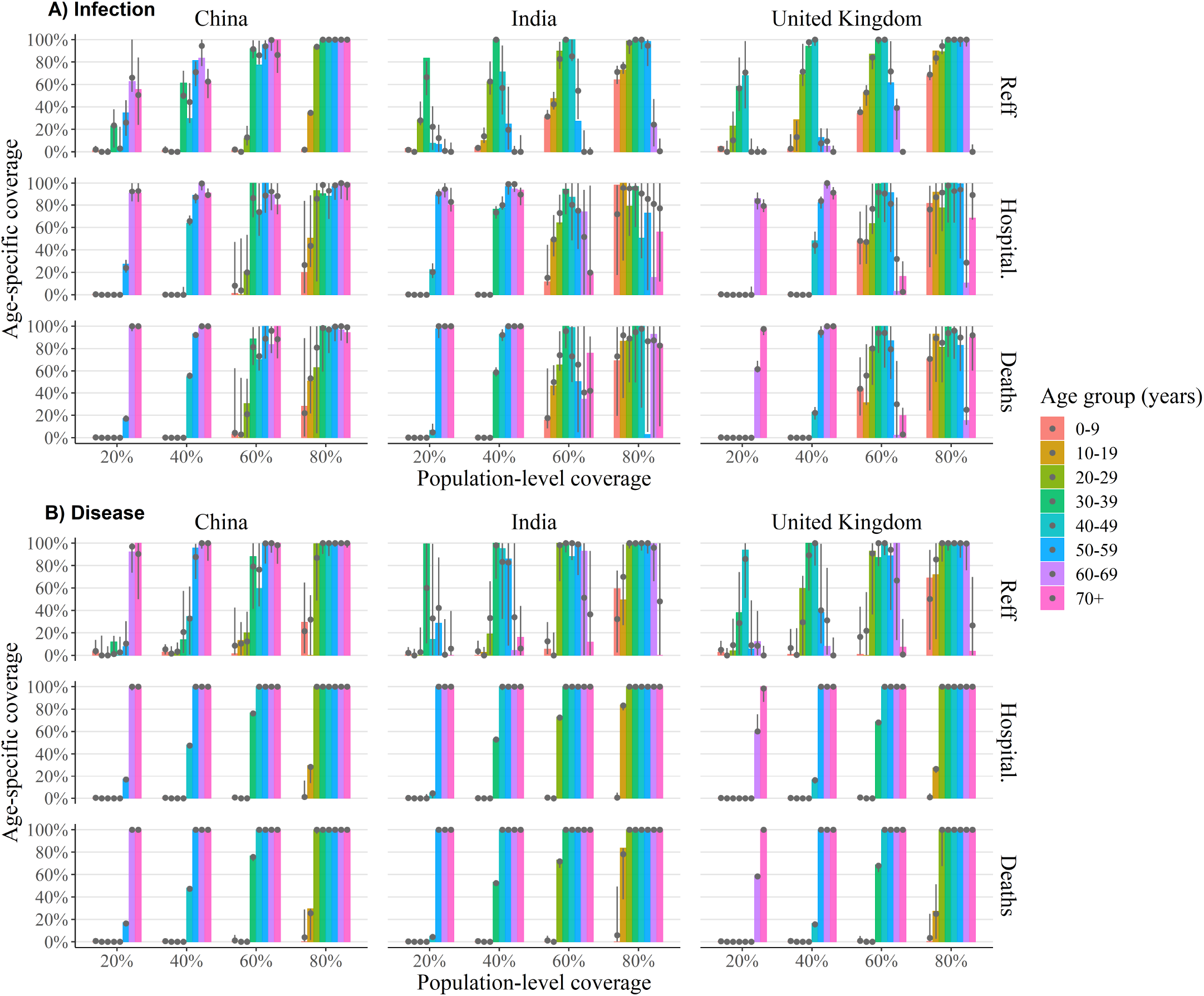
Optimal age-specific vaccination policy. The optimum coverage level for each age group (coloured bars) for infection-preventing (**A**) and disease-preventing (**B**) vaccines for varying population coverage levels to minimize transmission (top), hospitalizations (middle) and deaths (bottom) for India (left), China (middle) and the United Kingdom (right). Alongside the global optimal solution (coloured bars) we provide the 2.5 − 97.5 percentile ranges (vertical grey lines) and median values (grey dots) of age-specific coverage levels for all locally optimal solutions whose target value is within 1% of the global optimal value. Here we have assumed that the vaccine is 90% effective across all age groups (remaining model parameters assume their baseline values which can be found in Table S1).

In sensitivity analysis with reduced relative transmissibility among asymptomatic carriers, we find that disease-preventing vaccines become considerably more effective; however herd immunity remains unachievable in most settings (Figs. S5-S7). Alternatively, with reduced efficacy among 60+ year old individuals (i.e., 45% v. 90% in those <60 years old), we find similar results to those presented above, with changes in priority assigned to older individuals typically within the range of variation obtained from near-optimal solutions (grey bars) (Fig. S1). However, when we use an alternate model parameterization that only allows age-dependent variation in the clinical fraction (keeping individual susceptibility fixed across all age groups) we observe a shift in priority towards younger age groups — particularly in India and the UK (Figs. S9) — and a slight shift towards older age groups when we allow age-specific susceptibility only (with fixed clinical fraction by age) (Fig. S10).

### Efficiently reaching the herd immunity threshold

Following our analysis of optimal age-specific vaccine allocation under the constraint of finite available doses, we next investigated the inverse problem: calculating the minimum number of doses required to achieve herd immunity under targeted vaccine allocation. Given the results of the previous section indicating that vaccines that protect against symptomatic disease are often unable to achieve herd immunity, in this section we focused specifically on vaccines that reduce susceptibility to infection, which were shown to be more effective.

Under uniform vaccine allocation programs, we observe that, with a vaccine that is 90% effective at reducing susceptibility to infection, most countries require population-level vaccination coverages in excess of 60% (Fig. 4; top panel). In particular, we find that whilst vaccination appears to be a viable strategy to achieve herd immunity in some settings, more than 20% of the countries considered would require substantial coverage levels (>65%) to reach this goal.

**Figure 4.**
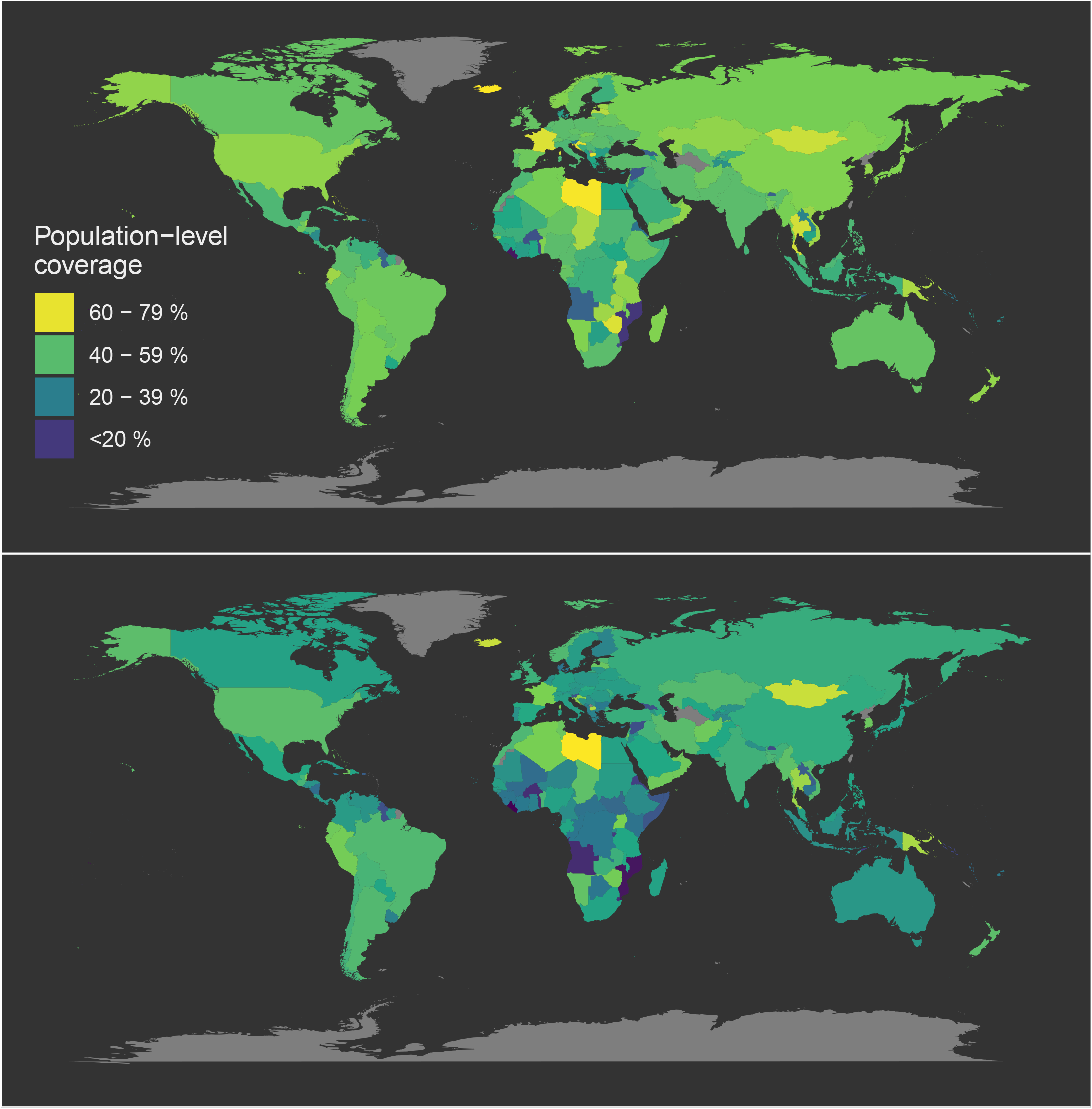
Global target vaccination coverage. Map of the minimum target vaccination coverage required to achieve herd immunity under country-specific uniform (top) and optimized (bottom) vaccine allocation programs.

In contrast, when dose allocation is optimized by targeting specific age groups, we observe dramatic reductions in the minimum population-level coverage required to achieve herd immunity (Fig. 4) — with most countries requiring less than 45% nationwide coverage under optimized, infection-preventing vaccination programs. As above, we find that countries with more intensely concentrated contacts within particular age groups experience greater (relative) reductions in the minimum population coverage required to achieve herd immunity under targeted vaccination programs, which in some cases can exceed 50% (e.g., Angola).

In line with baseline *R*_0_ estimates, the greatest variation in minimum coverage is observed across the African continent, which contains countries that require very low levels of vaccination (e.g., Angola, Mozambique), along with countries for which the minimum vaccination coverage required to achieve herd immunity may be unreachable (e.g., Libya at approximately 75%). Results obtained through sensitivity analysis with reduced vaccine efficacy in the 60+ year age groups (i.e., 45% v. 90% in those less than 60 years of age) are in most cases within a few percent of those presented in Fig. 4 (see Fig. S2). One notable exception is China, which under constant vaccine efficacy required only 48% (68%) population-level coverage under a targeted (uniform) vaccination policy, but is unable to reach the herd immunity threshold when efficacy wanes in 60+ year old individuals because of the high number of contacts amongst individuals in this age group.

We note that the minimum target values presented in Fig. 4 are highly sensitive to: (i) case-count-derived estimates of the basic reproduction numbers provided by Abbott et al.^16^ (and are therefore subject to biases introduced by variations in country-specific reporting rates through time); and (ii) the baseline vaccine efficacy (assumed here to be 90%). For comparison, we present the equivalent results for a vaccine that is 70% effective in Fig. S4, which are considerably higher — with several countries (e.g., France) failing to achieve herd immunity even with 100% population coverage. In general, our minimum coverage results can be readily rescaled without the need for re-optimization should updated estimates of *R*_0_ become available.

### Accounting for pre-existing immunity

To account for how substantial levels of infection-induced immunity prior to the introduction of vaccination modify the optimal age-specific dose allocation for each optimization target, we re-ran our optimization algorithm assuming a nationwide seropositivity of 25% in each of our three example countries (Fig. 5) (where in the United Kingdom the age-specific infection rates were extrapolated from a recent seroprevalence survey; see Methods). The only observable effect of pre-existing immunity is that for infection-preventing vaccines it allows an earlier switch towards an elimination strategy (i.e., prioritizing the middle-aged over the elderly) if minimizing morbidity and mortality are the primary objectives (Fig. 5, middle and bottom rows). Otherwise, the optimal age-specific vaccination coverages for minimizing transmission with infection-preventing vaccines, and minimizing all optimization targets for disease-preventing vaccines remained unaltered.

**Figure 5.**
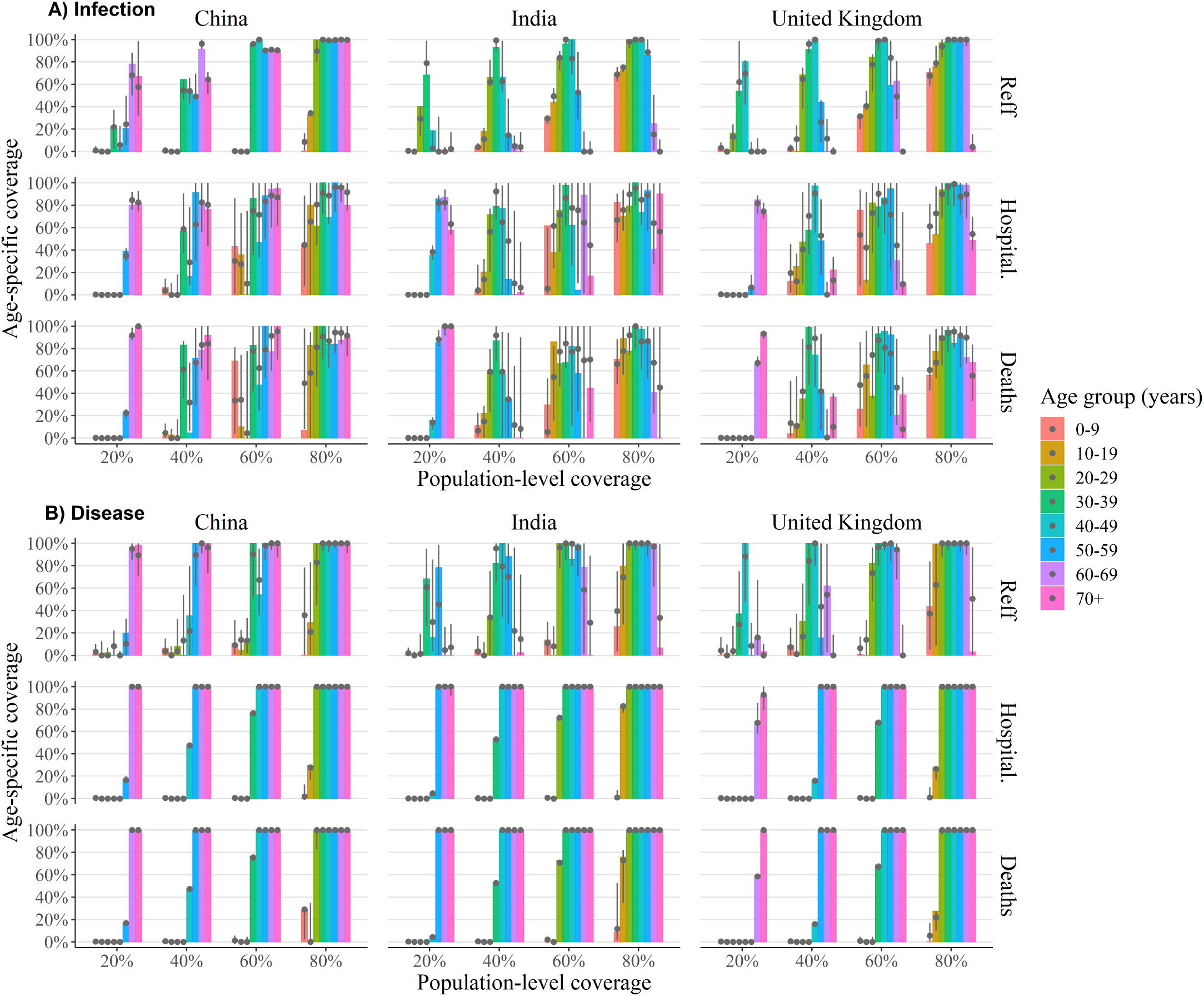
Optimal age-specific vaccination in the presence of pre-existing immunity. The optimum coverage level for each age group (coloured bars) for infection-preventing (**A**) and disease-preventing (**B**) vaccines for varying population coverage levels to minimize transmission (top), hospitalizations (middle) and deaths (bottom) in the United Kingdom when 25% of the population have infection-induced immunity. Alongside the global optimal solution (coloured bars) we provide the 2.5 − 97.5 percentile ranges (vertical grey lines) and median values (grey dots) of age-specific coverage levels for all locally optimal solutions whose target value is within 1% of the global optimal value. Here we have assumed that the vaccine is 90% effective for individuals aged 60 years and less and is 35% effective in those over the age of 60 (remaining model parameters assume their baseline values, which can be found in Tables S1 and S2).

## Discussion

Targeting vaccination towards age groups that contribute more to the transmission of infection, or are more likely to suffer severe outcomes following infection can dramatically enhance the effectiveness of COVID-19 vaccination. Whether the goal is to minimize transmission, morbidity or mortality for a fixed number of doses or to minimize the number of doses required to achieve a certain reduction in transmission, our analysis indicates that dosage requirements can be halved under tailored age-specific vaccination strategies. Whilst the choice of which age groups to initially target for vaccination is dependent on setting, vaccinating middle-aged individuals (30-59 years of age) typically had the highest impact on transmission across the 179 countries considered (e.g., India and the UK). However, there were some settings (e.g., China) in which those aged 60 and above were the first to be selected for vaccination (in line with observed contact structures^14^). Alternatively, prioritizing elderly individuals (60+ years of age) was the most efficient way to minimize hospitalizations and deaths, particularly when doses are limited. However, in general, when population-level vaccination coverage is sufficient to achieve herd immunity, we found that switching to a vaccination strategy that targets middle-aged individuals was optimal. This finding is supported by the recent analyses by^18,21,22^.

Additionally, the ability of vaccination to reduce transmission is highly dependent on the type of protection conferred^18^. Since asymptomatic individuals contribute substantially to SARS-CoV-2 transmission, vaccines that prevent symptomatic disease but allow transmissible asymptomatic infection failed to achieve herd immunity in the overwhelming majority of settings considered (assuming a baseline vaccine efficacy of 90%, in line with the recent announcements by Pfizer and Moderna). Conversely, vaccines that prevent initial infection by reducing the recipient’s susceptibility were far more likely to achieve elimination (and concomitantly possess greater scope for optimization). In either case, we emphasize that a vaccine with less than 100% efficacy may require substantial population-level coverage in order to achieve herd immunity^19^, although this threshold can be considerably reduced through targeted vaccination — as suggested by studies that account for heterogeneities in population susceptibility and mixing^11^.

Several serological surveys have revealed that pre-exisiting immunity generated by initial COVID-19 epidemic waves may have already reached a sizeable fraction of the optimal targets identified in our analysis^17,20^. Incorporating infection-induced immunity into our optimization algorithm, we found that the priority assigned to each age group remained largely unaltered for most combinations of optimization target and vaccination mode. The primary exceptions were minimizing hospitalizations and mortality with infection-preventing vaccines, where vaccinating middle-aged individuals (in an effort to achieve herd immunity) became optimal at lower values of population-level coverage.

Throughout our analysis we have attempted to account for parameter uncertainty by exploring a range of alternative vaccine profiles suggested by the World Health Organization’s recent vaccination modelling call^13^, as well as variations to our transmission modelling assumptions. Nevertheless, several limitations and possible extensions to the existing analysis remain. To begin, no vaccine provides perfect protection — and certainly not across all age groups. The results presented herein assume a vaccine efficacy of 90% that is sustained across all age groups at baseline; however, results assuming reduced efficacy (45%) in individuals over 60 years of age, and an alternative efficacy of 70% are presented in the Supplementary Materials. We found that reduced efficacy in individuals over 60 years of age had a negligible impact on the predicted minimum vaccination coverage targets and the level of priority assigned to particular age groups for most countries — China being a notable exception. Conversely, varying the baseline vaccine efficacy has a substantial impact on each of these outcomes: the minimum coverage required to achieve herd immunity increases substantially when the vaccine is 70% effective (with several countries unable to achieve herd immunity through vaccination alone — even with 100% coverage); similarly, the switch from elderly to middle-aged individuals as the highest priority age groups to minimize hospitalizations and mortality does not occur until higher values of population-level coverage are reached. Therefore, given that individual countries will have ranging access to an extensive suite of potential vaccines, it is important that the highly diverse efficacies of different candidates are considered when selecting an optimal strategy.

Next, when incorporating immunity as a result of past infection, we made the simplifying assumption that such immunity provides perfect protection against future infection: at this stage of the pandemic the true extent of protection provided by previous episodes of infection and disease over long time periods remains unclear; however, the exceedingly small number of cases that have reported repeat episodes suggests that infection-induced immunity is largely effective. Similarly, we assumed that vaccine doses are provided only to those individuals who remain susceptible, which in practice may require point-of-care testing to distinguish infection-naïve individuals from those that have already been infected and recovered. In the event such testing is unavailable, our results can be rescaled to account for “wasted” doses.

Furthermore, to avoid excessive speculation, we have not factored in the costs and feasibility of age-specific vaccine roll-out as part of our optimization search. However, alongside the global optimal solutions identified for each age group and setting, we have also indicated a range of possible age-specific coverage levels that have near-equivalent (within 1%) transmission reduction potential that may be selected if proven more cost-effective in practice.

In this modelling study, we have exclusively chosen to optimize vaccine allocation by age. This choice is motivated by the most readily available patient and contact data (which routinely provide breakdowns by age) and the subsequent analyses that have firmly established that infection and disease risk vary substantially by age. We anticipate that many vaccination programs will initially prioritize high-risk individuals (whose definition will undoubtedly be setting-specific) such as health- and aged-care workers. Therefore, if sufficient data is available, subsequent studies could also consider alternative demographic strata with known correlations with infection risk and disease severity, including occupation and socio-economic status, as well as the presence of comorbodities.

Another important limitation of our analysis is the reliance on case-based estimates for the basic reproduction number, *R*_0_. For each country, we have taken *R*_0_ to equal the maximum value of the time-varying *R*_eff_(*t*) estimate provided by the analysis in Abbott et al.^16^ (i.e., *R*_0_ = max_*t*_*R*_eff_(*t*)), and, to be conservative, we have used the upper 90% credible interval for *R*_eff_(*t*). Whilst variations in reporting rates through time within different host countries will likely bias these estimates, our predictions for the minimum vaccination coverage for each country can be re-scaled to account for any updated values.

We also note that the limited efficacy of disease-preventing vaccines reported in the main article stems from the prominence of sub-clinical transmitters, which, following Davies et al.^12^, we have assumed are 50% as infectious as clinical cases at baseline. As this parameter remains poorly quantified, we repeated our analysis for a range of relative transmissibilities and found that if the contribution of asymptomatic transmission (either in the natural infection case or post-vaccination) is lower, the estimated effectiveness of vaccines that prevent symptomatic disease, particularly for reducing transmission, correspondingly increases. Nevertheless, even when asymptomatic transmitters are only 25% as infectious as clinical cases, vaccines that prevent severe disease still fail to achieve herd immunity in many of the settings considered.

Another important consideration is the impact of waning vaccine-induced immunity, and the potentially harmful consequences of delaying an individual’s age-at-first-infection. This issue is known to influence the course of vaccination for several other communicable diseases (e.g., varicella^23^ and rubella) and may have important ramifications for COVID-19 that should be explored as more data become available — particularly given the significantly poorer outcomes observed among older individuals.

Finally, if and when a COVID-19 vaccine is approved and made available globally, it is critical that doses are allocated in a manner that most efficiently curtails transmission, morbidity and mortality — as initial supplies will undoubtedly be limited. Our study shows that setting- and age-specific optimization of vaccine allocation can provide substantial improvements in efficiency over uniform vaccination programs, saving up to half of the available doses to achieve the same reduction in each optimization target considered. Moreover, in deriving the minimum coverage thresholds required to achieve herd immunity in 179 countries, our analysis also underscores the often overlooked and unsavory outcome that vaccination alone may not be enough to completely suppress COVID-19 (particularly if the vaccine fails to prevent initial infection). In this case, permanent control measures — which may include physical distancing or improved hygiene — may be required alongside vaccination.

## Methods

### Calibrating transmission and the next-generation matrix

To model the transmission of SARS-CoV-2 we stratify the population into 16 5-year age bands *i* ∈ {0 − 4, 5 − 9, 10 − 14,…, 70 − 74, 75+} and assume that individuals in age group *i* possess a relative susceptibility to infection *u*_*i*_. Once infected, an age-dependent fraction *y*_*i*_ go on to develop symptomatic (i.e., clinical) disease whilst the remaining (1 − *y*_*i*_) develop asymptomatic (i.e., sub-clinical) disease. We assume that individuals in the sub-clinical class are less infectious than those in the clinical class by a relative factor *f* (baseline value 0.5^12^, range: 0.25-0.75) and that the total time spent infectious for both classes is *τ* = 5.0 days^12^.

Each day, each individual in age group *j* makes *c*_*i j*_ contacts with individuals in age group *i* leading to the following expression for the (unscaled) next-generation matrix (NGM)^12,24^:

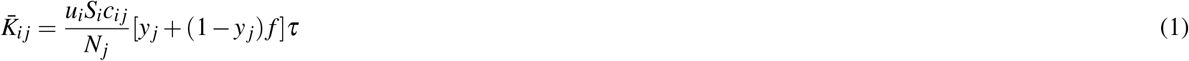

where *S*_*i*_ is the number of susceptible individuals in age group *i* and *N*_*j*_ is total number of individuals in age group *j*. We allow that prior to vaccination an age-specific fraction *p*_*i*_ of individuals have immunity as a result of previous infection, such that *S*_*i*_ = (1 − *p*_*i*_)*N*_*i*_.

The (*i, j*)th entry of the NGM 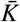 is proportional to the average number of new infections in age group *i* generated by an individual in age group *j* over their entire infectious lifetime. To calculate the actual number of infections generated by each individual these entries must be scaled by the (pseudo-)probability of transmission given contact, which we denote by *η*. In particular, the basic reproduction number, *R*_0_, which is proportional to the maximal eigenvalue of 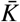, can be expressed as

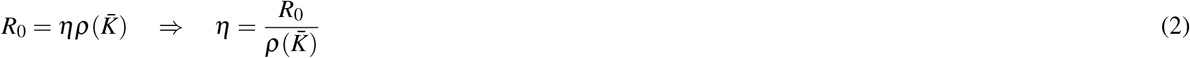

where we use 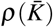 to denote the spectral radius of the matrix 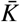.

Incorporating this definition of *η* along with the possibility of pre-existing immunity (*p*_*i*_) the next-generation matrix *K* is given by

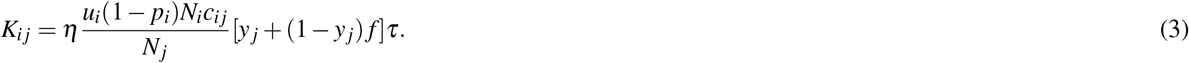

For each of the 179 countries considered in our analysis, we used estimates of *R*_0_ based on observed case counts^16^ to calibrate country-specific values for the scaling factor *η*. In particular, we assume *R*_0_ in each country is equal to the maximum value of the 90% credible interval upper limit of the time-varying reproduction number estimates generated by Abbott et al.^16^. Where these values are unrealistically high (potentially due to large variations in reporting rates through time; e.g., North Macedonia returned an *R*_0_ greater than 30), we used estimates provided by the Centres for Disease Control and Prevention (CDC).

We assume that the age-dependent susceptibility (*u*_*i*_) and clinical fraction (*y*_*i*_) (i.e., biological quantities) are universal across all countries and allow only the age-specific contact rates (*c*_*i j*_), initial population sizes (*S*_*i*_ = (1 − *p*_*i*_)*N*_*i*_ and *N*_*j*_) and transmission scaling factors *η* to vary among settings, i.e., all other parameters remain fixed among countries (see Table S1). We consider three separate models for age-dependent susceptibility and clinical fraction: M1 — allowing both age-dependent susceptibility and clinical fraction; M2 — allowing age-dependent susceptibility only; and M3 — allowing age-dependent clinical fraction only. The results for model M1 are those presented in the main article, whilst results for models M2 and M3 appear in the Supplement. Parameter values under each model have been calibrated to patient data in Davies et al.^12^ and can be found in Table S2.

Where available, the daily, age-dependent contact rates, *c*_*i j*_, between individuals in each country were taken from previously conducted nationwide contact surveys. This was the case for China^14^ and the UK^15^, as well Italy, Germany, Luxembourg, the Netherlands, Poland, Finland and Belgium^25^ and Zimbabwe^26^. In the absence of such survey data, synthetic contact matrices extrapolated from existing contact surveys across multiple settings were used^27,28^.

At baseline, each country is assumed to have negligible infection rates (relative to population size) such that *p*_*i*_ = 0. To test the robustness of our results to this assumption we re-ran our analyses with a baseline seroprevalence of 25%. For simplicity we assumed that past infection confers complete protection against future infection, and that available doses are provided exclusively to infection-naïve individuals. In the United Kingdom age-specific infection rates are calculated by upscaling the infection rates observed in a recent seroprevalence survey^17^ to achieve a nationwide seroprevalence of 25%, whilst in China and India (where currently reported seroprevalence estimates are considerably lower^29^) we assumed a uniform infection rate across all age groups.

### Calculating disease-related morbidity and mortality

To calculate the total number of hospitalized cases and deaths throughout the course of the epidemic we use the vectorized form of the final size equation given by^30^:

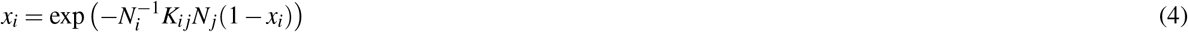

where *x*_*i*_ = *S*_*i*_(∞)*/S*_*i*_(0) is the fraction of individuals that remain uninfected at the completion of the epidemic, and *N*_*i*_ and *K*_*i j*_ are defined in the previous section. Solving the final size equation numerically, we can determine the total number of hospitalizations (*H*) and deaths (*D*) using the following expressions:

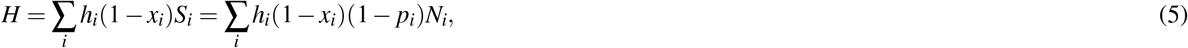

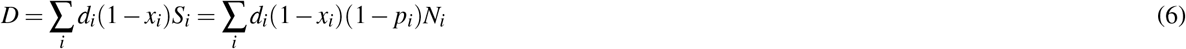

where *h*_*i*_ is the age-specific hospitalization rate among all infected individuals^31^ and *d*_*i*_ is the age-specific infection-fatality-rate^32^. These values are listed in Table S3.

### Modelling vaccination

To incorporate vaccination of otherwise susceptible individuals, we let *v*_*i*_ denote the fraction of individuals in age group *i* that are vaccinated (i.e., the coverage of age-group *i*), *e* denote the baseline efficacy of the vaccine, and *r*_*i*_ its age-dependent relative efficacy. For tractability, here we divide the population into 10-year age bands such that the vector *v*_*i*_ has eight free parameters that are repeated pairwise to cover the 16 5-year age bands defined above.

We investigate two distinct modes of action for potential vaccines, those that protect against initial infection and those that protect against symptomatic disease following infection. The former equates to a removal of susceptible individuals from the total population (similar to the role of infection-induced immunity) whilst the latter influences the fraction of individuals that go on to develop symptomatic disease. The NGM in each instance transforms, respectively, to:

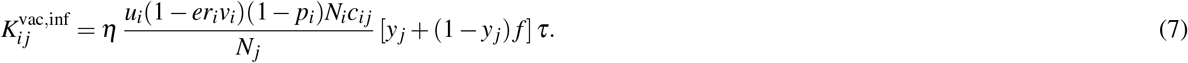

and

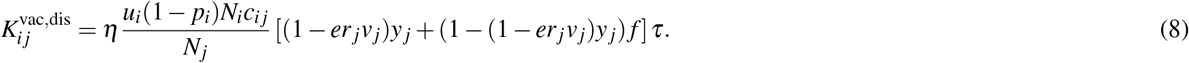

In the presence of vaccination, the effective reproduction number, *R*_eff_, can be calculated via

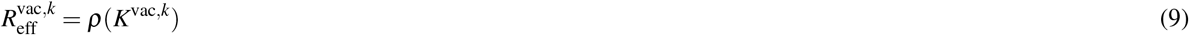

where the index *k* ∈ {inf, dis} denotes the type of protection conferred by vaccination, and the scaling factor *η* is calibrated according to the procedure described in the previous section.

Similarly, the total number of hospitalizations and deaths in the presence of vaccination are determined respectively by

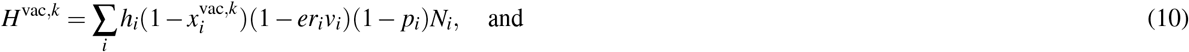

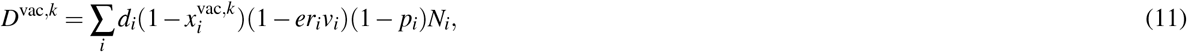

where *x*^vac,*k*^ is the solution to the vaccination-modified final size equation

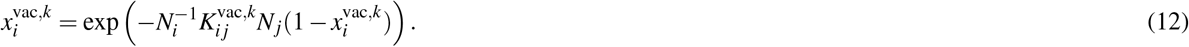

Note in the expressions for *H*^vac,dis^ and *D*^vac,dis^ given above, we have assumed that all COVID-related hospitalizations and deaths result from infected individuals who develop severe disease.

### Optimizing vaccination

Using expression (9) for the effective reproduction number, the total number of hospitalizations, and the total number of deaths as a function of the age-specific vaccination proportions **v** = (*v*_0−9_, *v*_10−19_, …, *v*_70+_) respectively, we pursue four optimization targets: 1) minimizing the effective reproduction number (*R*_eff_) for a given (fixed) number of doses; 2) minimizing the total number of hospitalizations (*H*) for a fixed number of doses; 3) minimizing the total number of deaths (*D*) for a fixed number of doses; and 4) minimizing the total number of doses (i.e., population-level coverage) required to achieve herd immunity, which we define as obtaining an effective reproduction number less than one. These optimization problems can respectively be stated as

1. **Minimizing transmission with a fixed number of doses**.

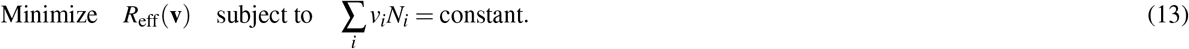
  2. **Minimizing hospitalizations with a fixed number of doses**.

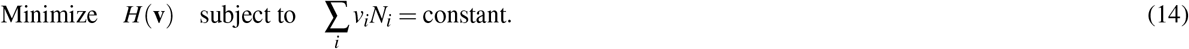
  2. **Minimizing deaths with a fixed number of doses**.

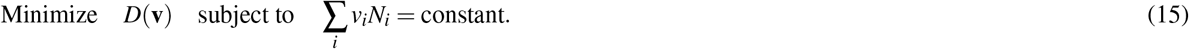
  4. **Efficiently reaching the herd immunity threshold**.

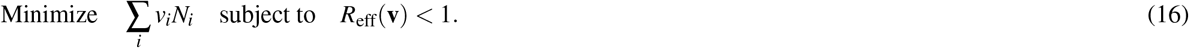

Since the surfaces *R*_eff_(**v**), *H*(**v**), *D*(**v**) = const. are highly complex, we expect that there may be multiple solutions to the optimization problems 1 - 4. Therefore, we used a stochastic simulated annealing algorithm to search the parameter space and re-ran it multiple times to uncover multiple solutions of roughly equivalent quality (i.e., similar objective values). Hence, in addition to storing the global optimum value (i.e., the solution with the minimum objective value among all solutions obtained) we also retained the values obtained from each of the remaining runs whose objective value was within 1% of the global optimum (the range of these values is indicated by the grey bars in the age-specific vaccination coverage figures, e.g., Fig. 3).

## Data Availability

All input data and model source code has been deposited in a recognized public source repository (GitHub, https://github.com/michaeltmeehan/covid19).

## Data and code availability

All input data and model source code has been deposited in a recognized public source repository GitHub, https://github.com/michaeltmeehan/covid19. Source code has been written in the julia programming language.

## Acknowledgements

M.M. is funded by a postdoctoral research fellowship from the National Health and Medical Research Council’s Centre of Research Excellence in Tuberculosis Control on Both Sides of Our Border (Project ID: 1153493).

## Author contributions statement

M.M., R.R and E.M. conceived the project, M.M. and D.C. conducted the simulations, M.M., J.C., D.C., A.A., J.T., R.R. and E.M. analysed the results. All authors reviewed the manuscript.

## Additional information

### Competing interests

The authors do not have any competing interests to declare.

## Supplementary Information

In addition to the data provided below, further model inputs and results — including optimization results for all 179 countries — can be found in the public GitHub repository: https://github.com/michaeltmeehan/covid19/optimize_vaccination.

**Table S1.**
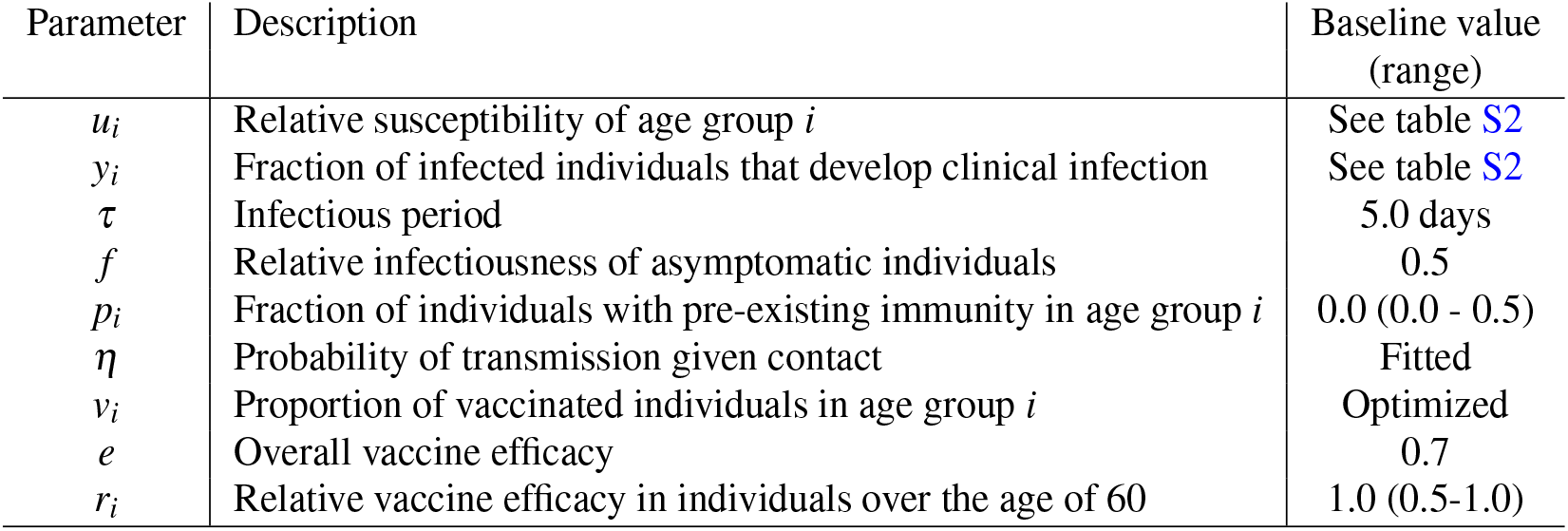
Transmission model parameters adapted from^12^.

**Table S2.** Fitted values of age-dependent susceptibility and clinical fraction generated by Davies et al.^12^ allowing for both age-dependent susceptibility and clinical fraction (M1); age-dependent susceptibility only (M2); and age-dependent clinical fraction only (M3).

| Age-group (years) | Relative susceptibility ($u_i$) | | Clinical fraction ($y_i$) | |
| --- | --- | --- | --- | --- |
|  | M1 | (M2, M3) | M1 | (M2, M3) |
| 0 – 4 | 0.39 | (0.018, 1.0) | 0.28 | (0.50, 0.14) |
| 5 – 9 | 0.39 | (0.018, 1.0) | 0.28 | (0.50, 0.14) |
| 10 – 14 | 0.38 | (0.018, 1.0) | 0.20 | (0.50, 0.14) |
| 15 – 19 | 0.38 | (0.018, 1.0) | 0.20 | (0.50, 0.15) |
| 20 – 24 | 0.79 | (0.022, 1.0) | 0.26 | (0.50, 0.18) |
| 25 – 29 | 0.79 | (0.028, 1.0) | 0.26 | (0.50, 0.23) |
| 30 – 34 | 0.87 | (0.036, 1.0) | 0.33 | (0.50, 0.29) |
| 35 – 39 | 0.87 | (0.042, 1.0) | 0.33 | (0.50, 0.34) |
| 40 – 44 | 0.80 | (0.046, 1.0) | 0.40 | (0.50, 0.38) |
| 45 – 49 | 0.80 | (0.049, 1.0) | 0.40 | (0.50, 0.40) |
| 50 – 54 | 0.82 | (0.053, 1.0) | 0.49 | (0.50., 0.44) |
| 55 – 59 | 0.82 | (0.060, 1.0) | 0.49 | (0.50, 0.50) |
| 60 – 64 | 0.89 | (0.067, 1.0) | 0.63 | (0.50, 0.55) |
| 65 – 69 | 0.89 | (0.070, 1.0) | 0.63 | (0.50, 0.57) |
| 70 – 74 | 0.74 | (0.071, 1.0) | 0.69 | (0.50, 0.58) |
| 75+ | 0.74 | (0.071, 1.0) | 0.69 | (0.50, 0.58) |

**Table S3.** Age-specific hospitalization^31^ (*h*_*i*_) and fatality^32^ (*d*_*i*_) rates among all infected individuals.

| Age-group (years) | Infection hospitalization rate (%) ($h_i$) | Infection fatality rate (%) ($d_i$) |
| --- | --- | --- |
| 0 – 4 | 0.10 | 0.0030 |
| 5 – 9 | 0.10 | 0.00069 |
| 10 – 14 | 0.10 | 0.0011 |
| 15 – 19 | 0.20 | 0.0026 |
| 20 – 24 | 0.50 | 0.0079 |
| 25 – 29 | 1.0 | 0.017 |
| 30 – 34 | 1.6 | 0.033 |
| 35 – 39 | 2.3 | 0.055 |
| 40 – 44 | 2.9 | 0.11 |
| 45 – 49 | 3.9 | 0.17 |
| 50 – 54 | 5.8 | 0.30 |
| 55 – 59 | 7.2 | 0.46 |
| 60 – 64 | 10.2 | 0.60 |
| 65 – 69 | 11.7 | 1.5 |
| 70 – 74 | 14.6 | 2.4 |
| 75+ | 17.7 | 4.3 |

**Figure S1.**
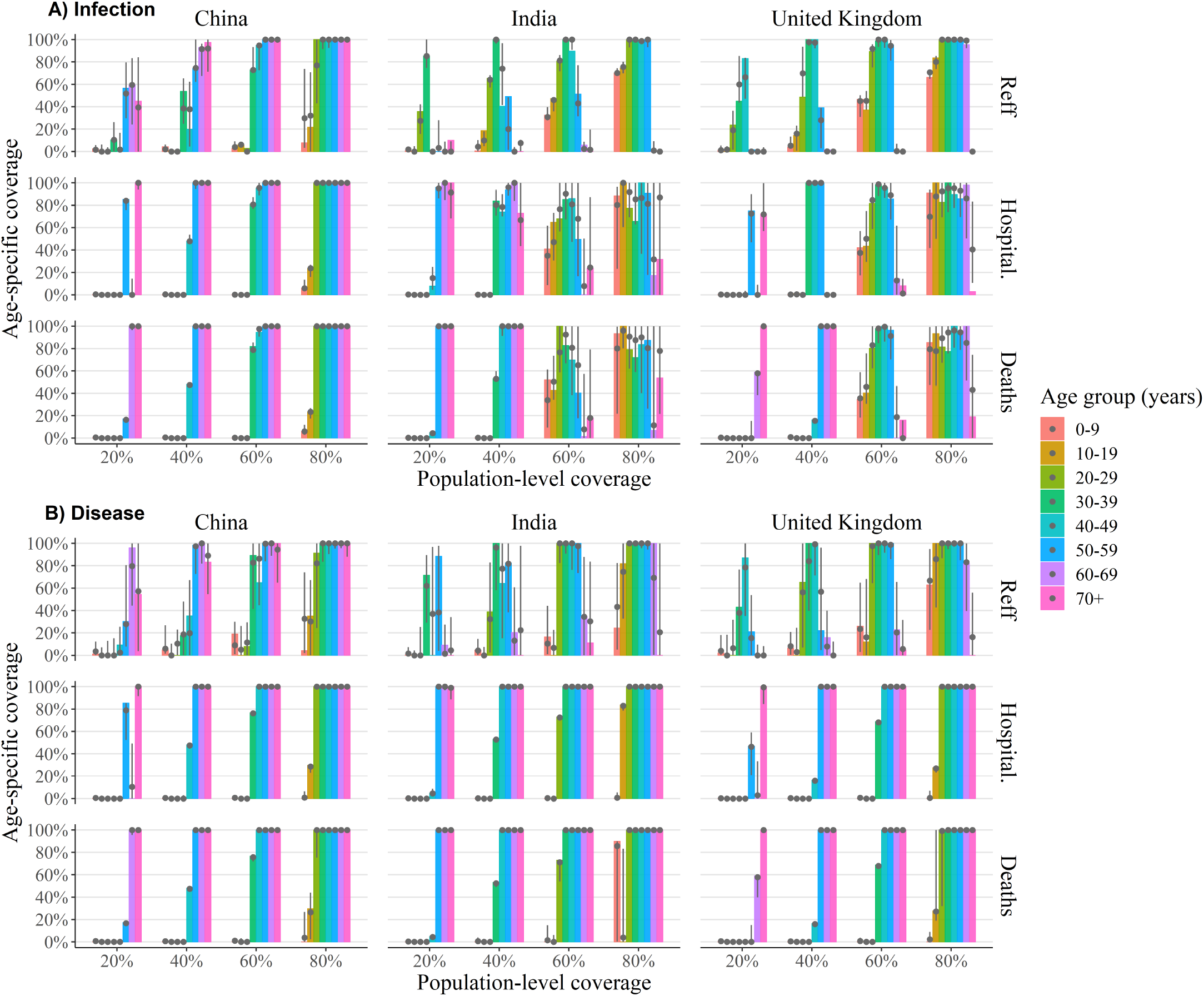
Optimal age-specific vaccination policy (with reduced efficacy in 60+ year olds). The optimum coverage level for each age group (coloured bars) for infection-preventing (**A**) and disease-preventing (**B**) vaccines for varying population coverage levels to minimize transmission (top), hospitalizations (middle) and deaths (bottom) for India (left), China (middle) and the United Kingdom (right). Alongside the global optimal solution (coloured bars) we provide the 2.5 − 97.5 percentile ranges (vertical grey lines) and median values (grey dots) of age-specific coverage levels for all locally optimal solutions whose target value is within 1% of the global optimal value. Here we have assumed that the vaccine is 90% effective for individuals aged 60 years and less and is 35% effective in those over the age of 60 (remaining model parameters assume their baseline values, which can be found in Tables S1 and S2).

**Figure S2.**
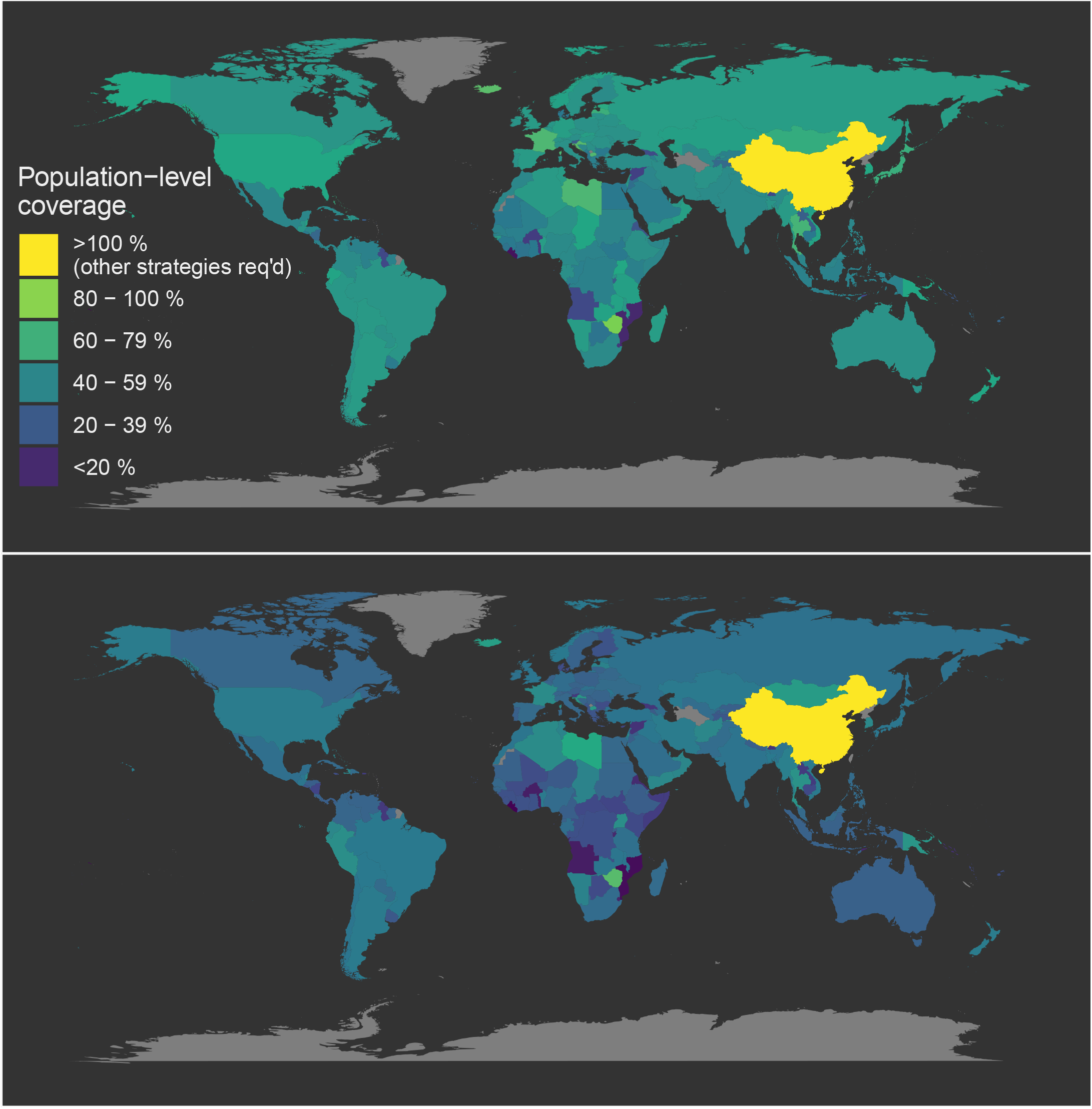
Global target vaccination coverage under uniform vaccination policy (with reduced efficacy in 60+ year olds). Map of the minimum target vaccination coverage required to achieve herd immunity under uniform vaccine allocation programs. Countries coloured bright yellow are incapable of achieving herd immunity through vaccination alone (i.e., their minimum coverage thresholds exceed 100%). Here we have assumed that the vaccine is 70% effective for individuals aged 60 years and less and is 35% effective in those over the age of 60 (remaining model parameters assume their baseline values, which can be found in Tables S1 and S2).

**Figure S3.**
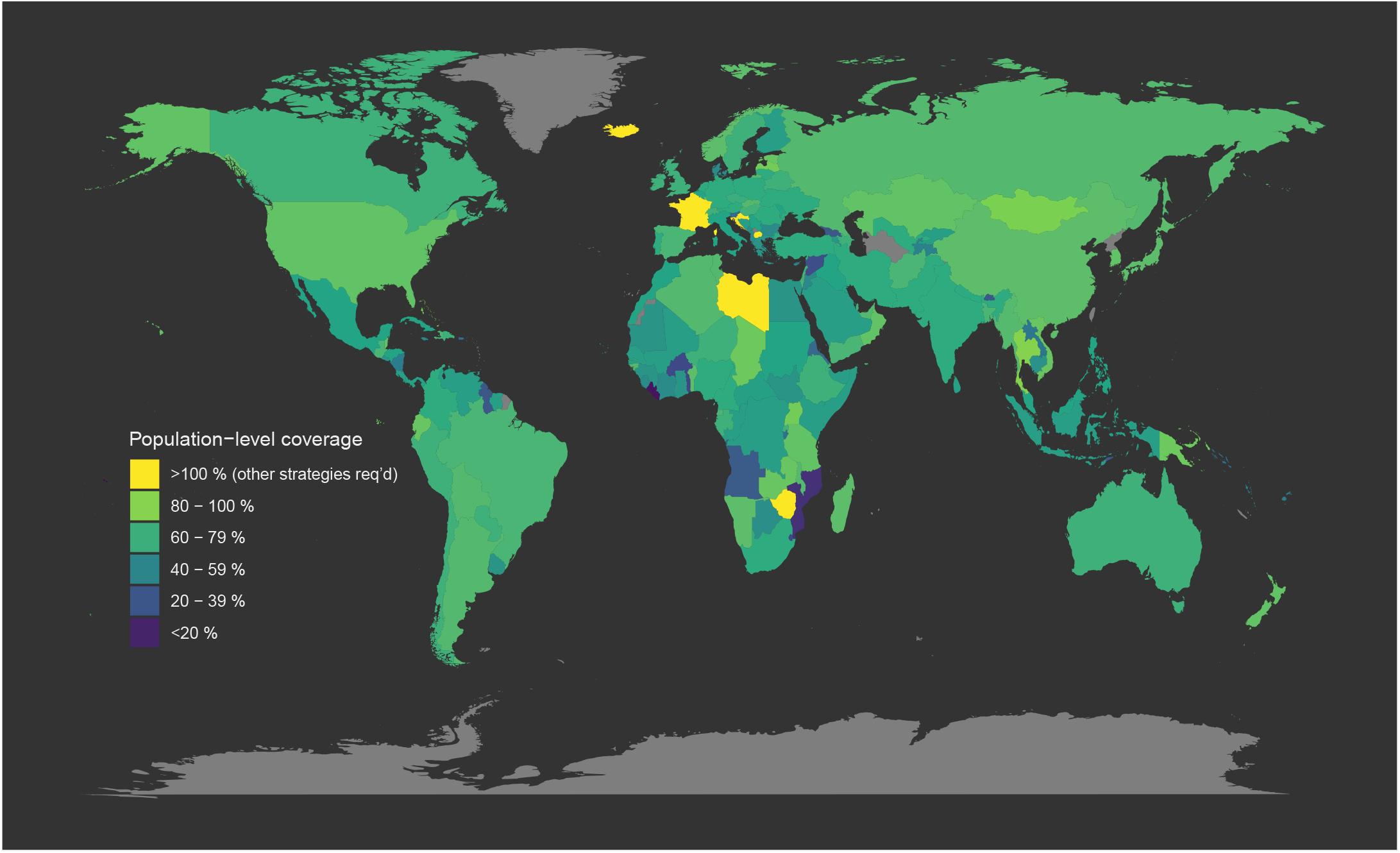
Global target vaccination coverage under uniform vaccination policy (with a baseline vaccine efficacy of 70%). Map of the minimum target vaccination coverage required to achieve herd immunity under uniform vaccine allocation programs. Countries coloured bright yellow are incapable of achieving herd immunity through vaccination alone (i.e., their minimum coverage thresholds exceed 100%).

**Figure S4.**
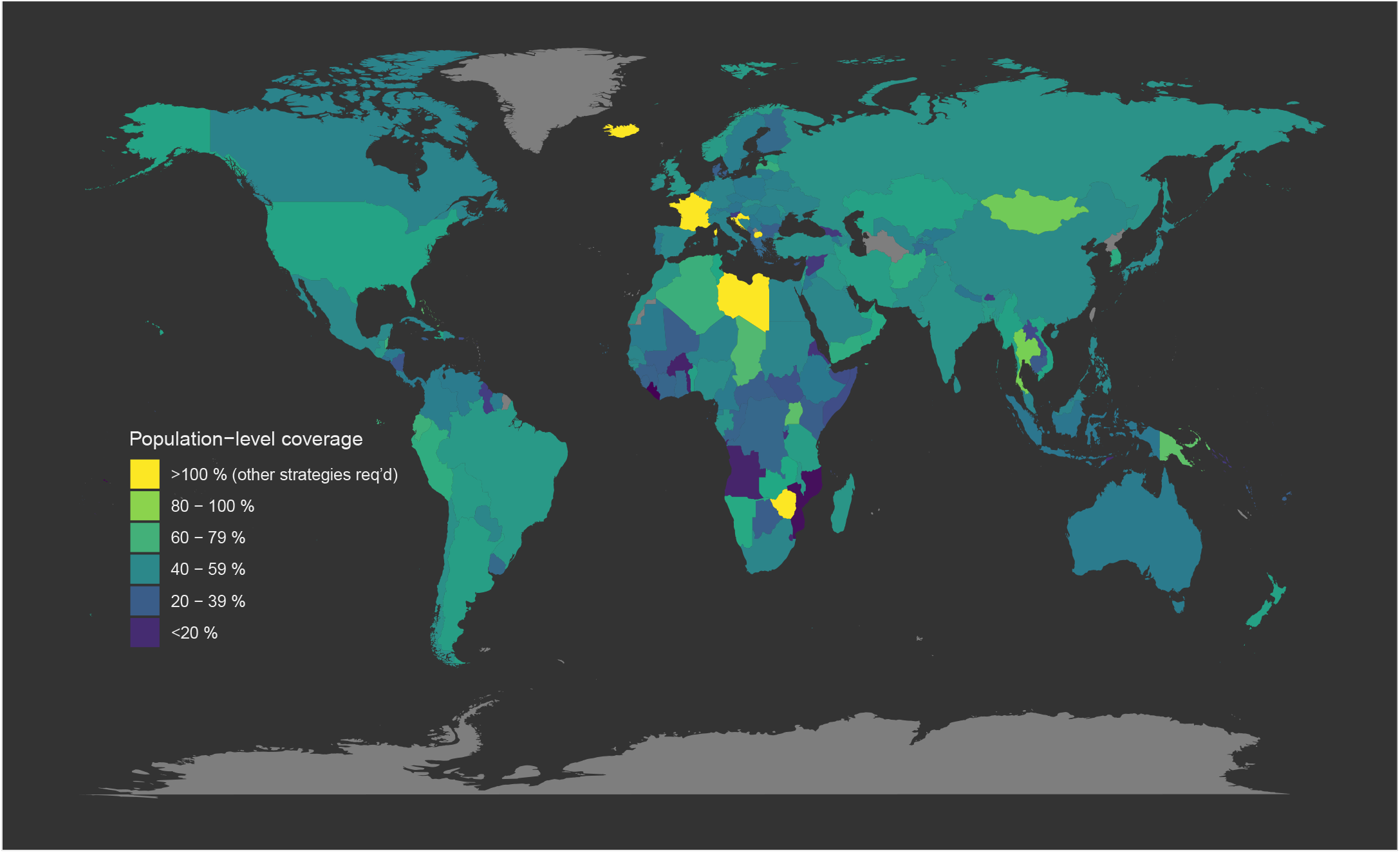
Global target vaccination coverage under optimized vaccination policy (with a baseline vaccine efficacy of 70%). Map of the minimum target vaccination coverage required to achieve herd immunity under country-specific optimized vaccine allocation programs. Countries coloured yellow are incapable of achieving herd immunity through vaccination alone (i.e., their minimum coverage threshold exceeds 100%).

**Figure S5.**
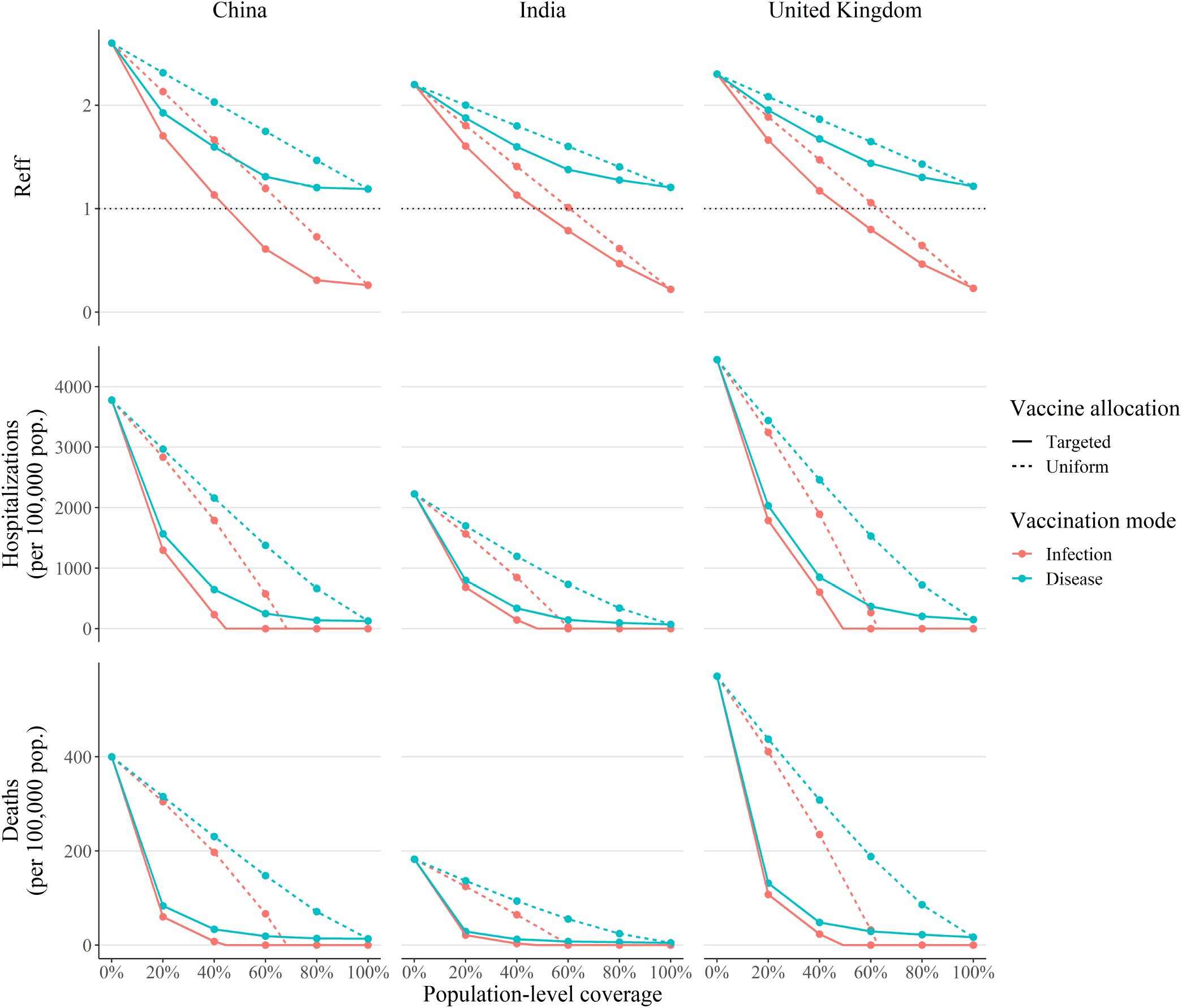
Reduction in transmission, hospitalizations and deaths under optimized vaccine allocation (with asymptomatic individuals 25% as infectious as clinical cases). Effective reproduction number *R*_eff_ (top), cumulative hospitalizations (middle) and deaths (bottom), as a function of the population-level vaccination coverage when allocation is targeted towards priority age groups (solid lines) and the vaccine protects against infection (red) and disease (blue) for India (left), China (middle) and the United Kingdom (right). For comparison, we also plot the analogous reduction in the three optimization targets when vaccine doses are allocated uniformly across age groups (dashed lines). In the top panel row, the herd immunity threshold (*R*_eff_<1) is indicated by the horizontal dotted black line.

**Figure S6.**
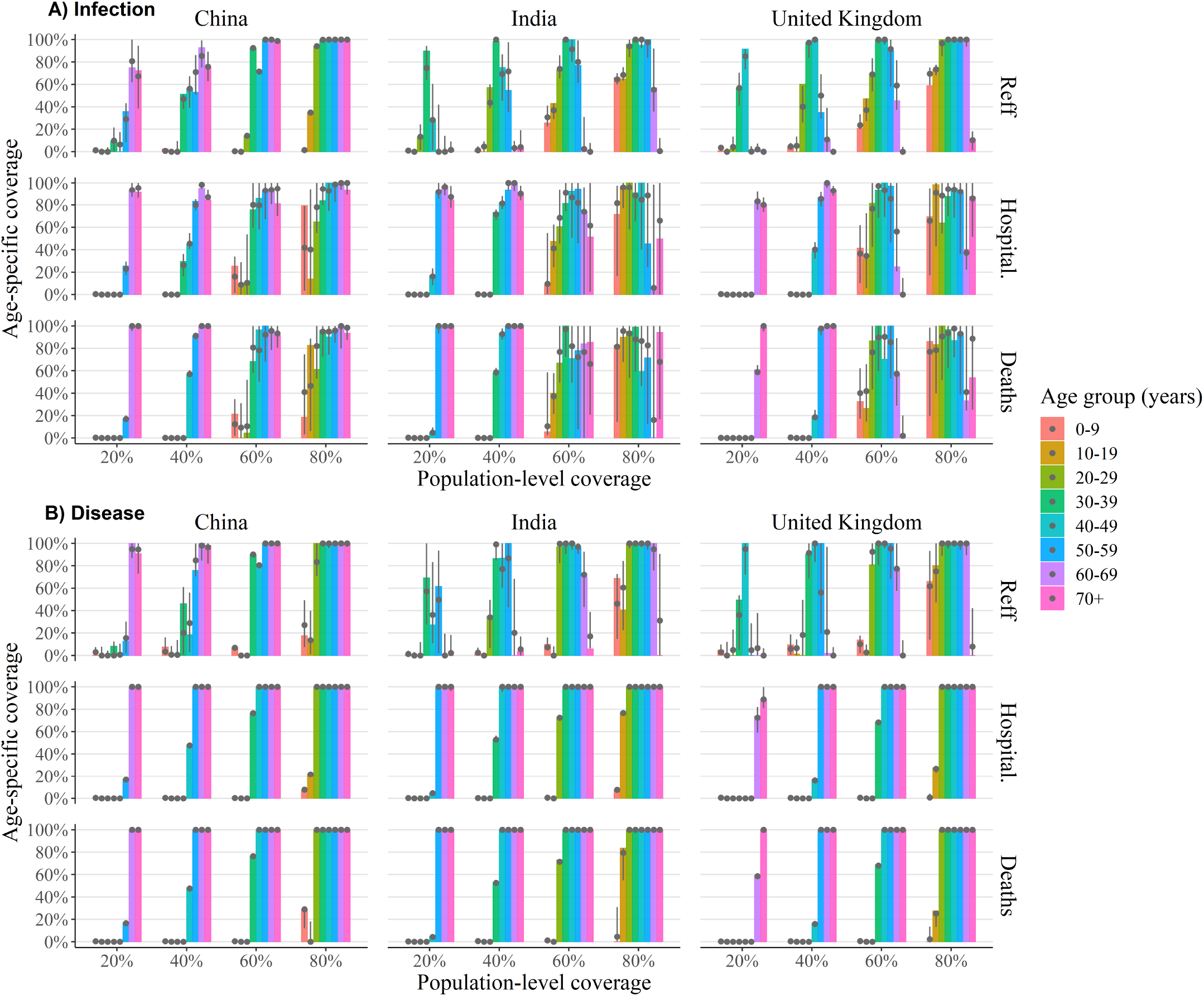
Optimal age-specific vaccination policy (with asymptomatic individuals 25% as infectious as clinical cases). The optimum coverage level for each age group (coloured bars) for infection-preventing (**A**) and disease-preventing (**B**) vaccines for varying population coverage levels to minimize transmission (top), hospitalizations (middle) and deaths (bottom) for India (left), China (middle) and the United Kingdom (right). Alongside the global optimal solution (coloured bars) we provide the 2.5 − 97.5 percentile ranges (vertical grey lines) and median values (grey dots) of age-specific coverage levels for all locally optimal solutions whose target value is within 1% of the global optimal value. Here we have assumed that the vaccine is 90% effective across all age groups (remaining model parameters assume their baseline values which can be found in Table S1).

**Figure S7.**
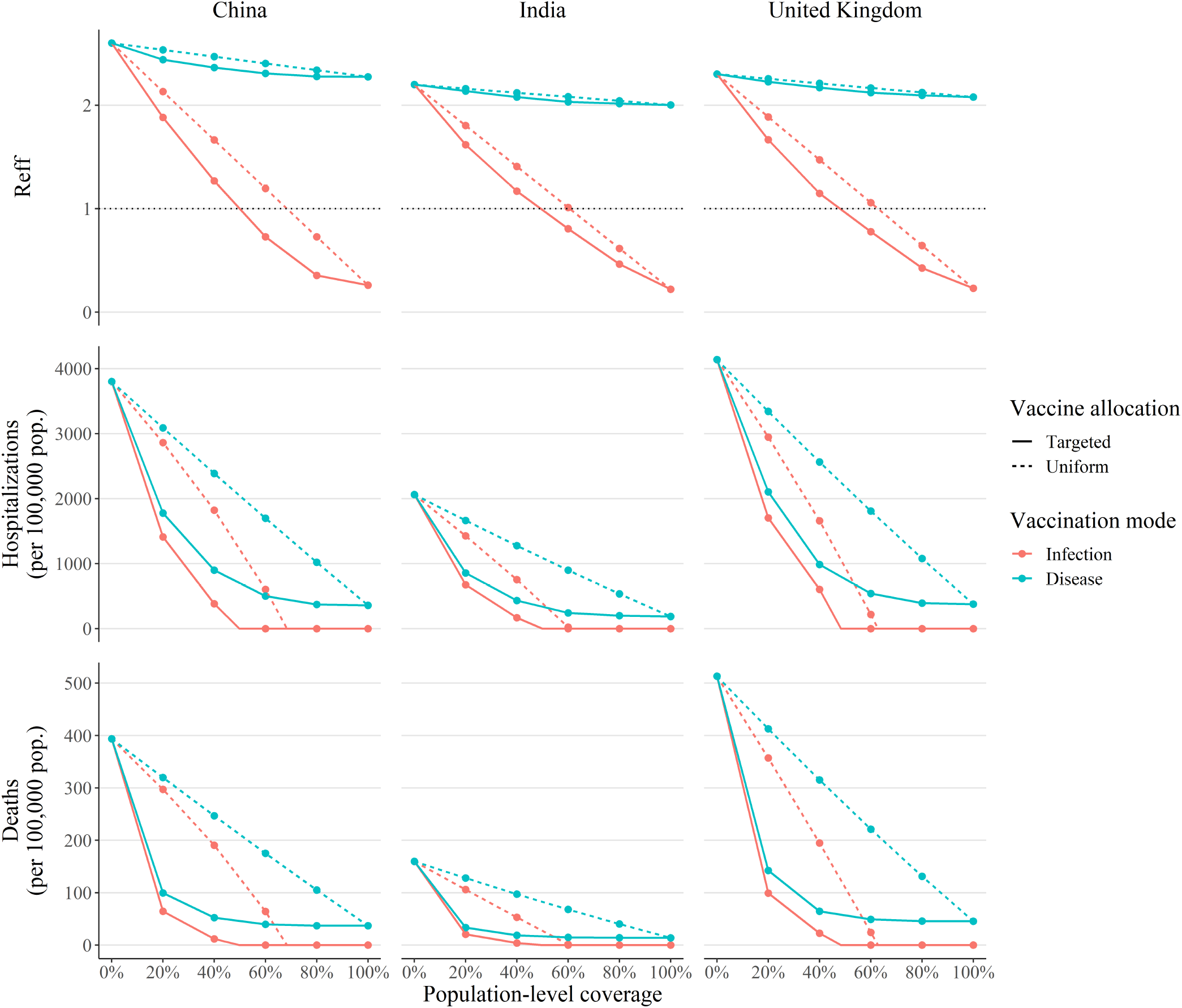
Reduction in transmission, hospitalizations and deaths under optimized vaccine allocation (with asymptomatic individuals 75% as infectious as clinical cases). Effective reproduction number *R*_eff_ (top), cumulative hospitalizations (middle) and deaths (bottom), as a function of the population-level vaccination coverage when allocation is targeted towards priority age groups (solid lines) and the vaccine protects against infection (red) and disease (blue) for India (left), China (middle) and the United Kingdom (right). For comparison, we also plot the analogous reduction in the three optimization targets when vaccine doses are allocated uniformly across age groups (dashed lines). In the top panel row, the herd immunity threshold (*R*_eff_<1) is indicated by the horizontal dotted black line.

**Figure S8.**
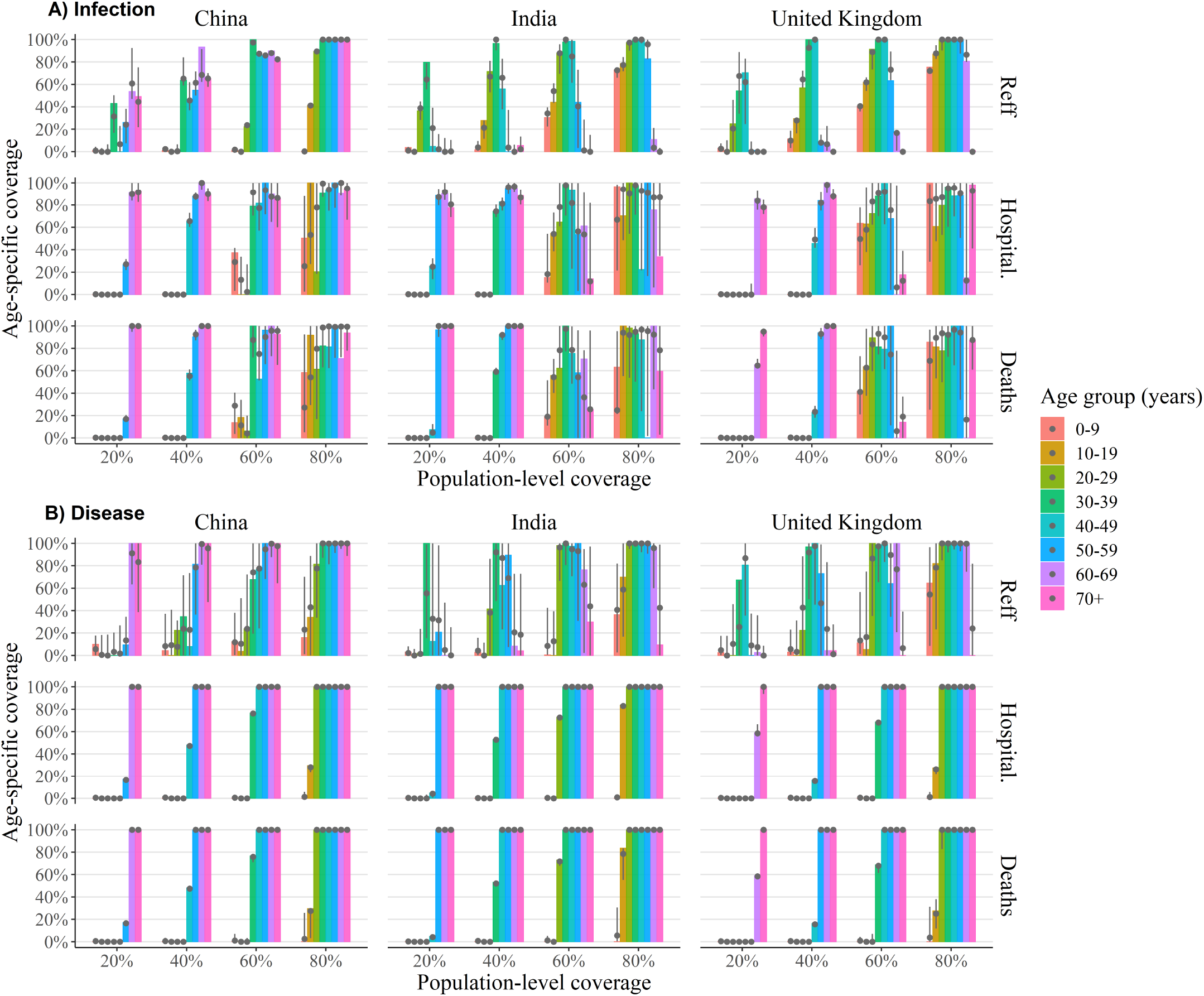
Optimal age-specific vaccination policy (with asymptomatic individuals 75% as infectious as clinical cases). The optimum coverage level for each age group (coloured bars) for infection-preventing (**A**) and disease-preventing (**B**) vaccines for varying population coverage levels to minimize transmission (top), hospitalizations (middle) and deaths (bottom) for India (left), China (middle) and the United Kingdom (right). Alongside the global optimal solution (coloured bars) we provide the 2.5 − 97.5 percentile ranges (vertical grey lines) and median values (grey dots) of age-specific coverage levels for all locally optimal solutions whose target value is within 1% of the global optimal value. Here we have assumed that the vaccine is 90% effective across all age groups (remaining model parameters assume their baseline values which can be found in Table S1).

**Figure S9.**
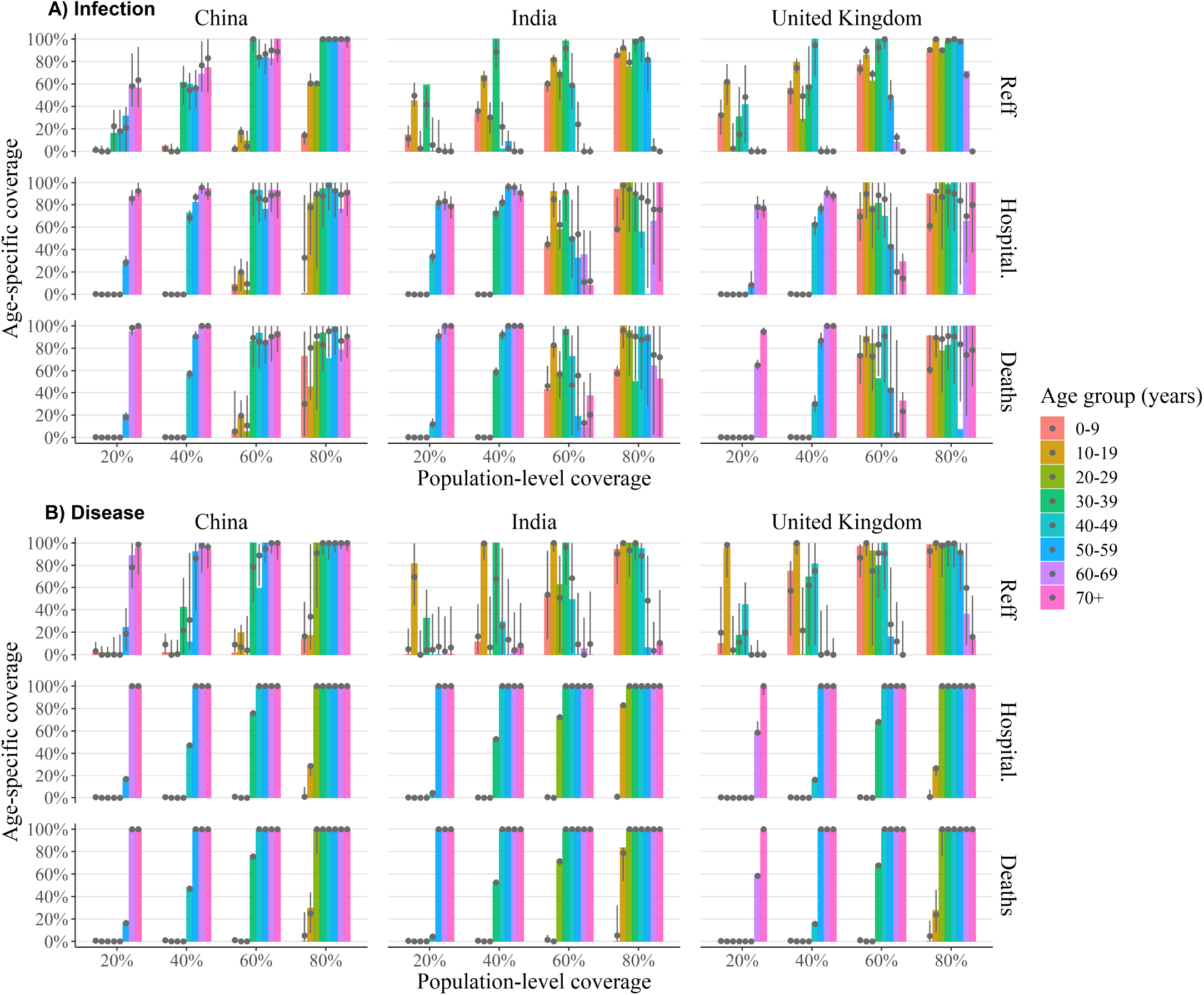
Optimal age-specific vaccination policy, allowing age-dependent susceptibility only (M2). The optimum coverage level for each age group (coloured bars) for infection-preventing (**A**) and disease-preventing (**B**) vaccines for varying population coverage levels to minimize transmission (top), hospitalizations (middle) and deaths (bottom) for India (left), China (middle) and the United Kingdom (right). Alongside the global optimal solution (coloured bars) we provide the 2.5 − 97.5 percentile ranges (vertical grey lines) and median values (grey dots) of age-specific coverage levels for all locally optimal solutions whose target value is within 1% of the global optimal value. Here we have assumed that the vaccine is 90% effective across all age groups (remaining model parameters assume their baseline values which can be found in Table S1).

**Figure S10.**
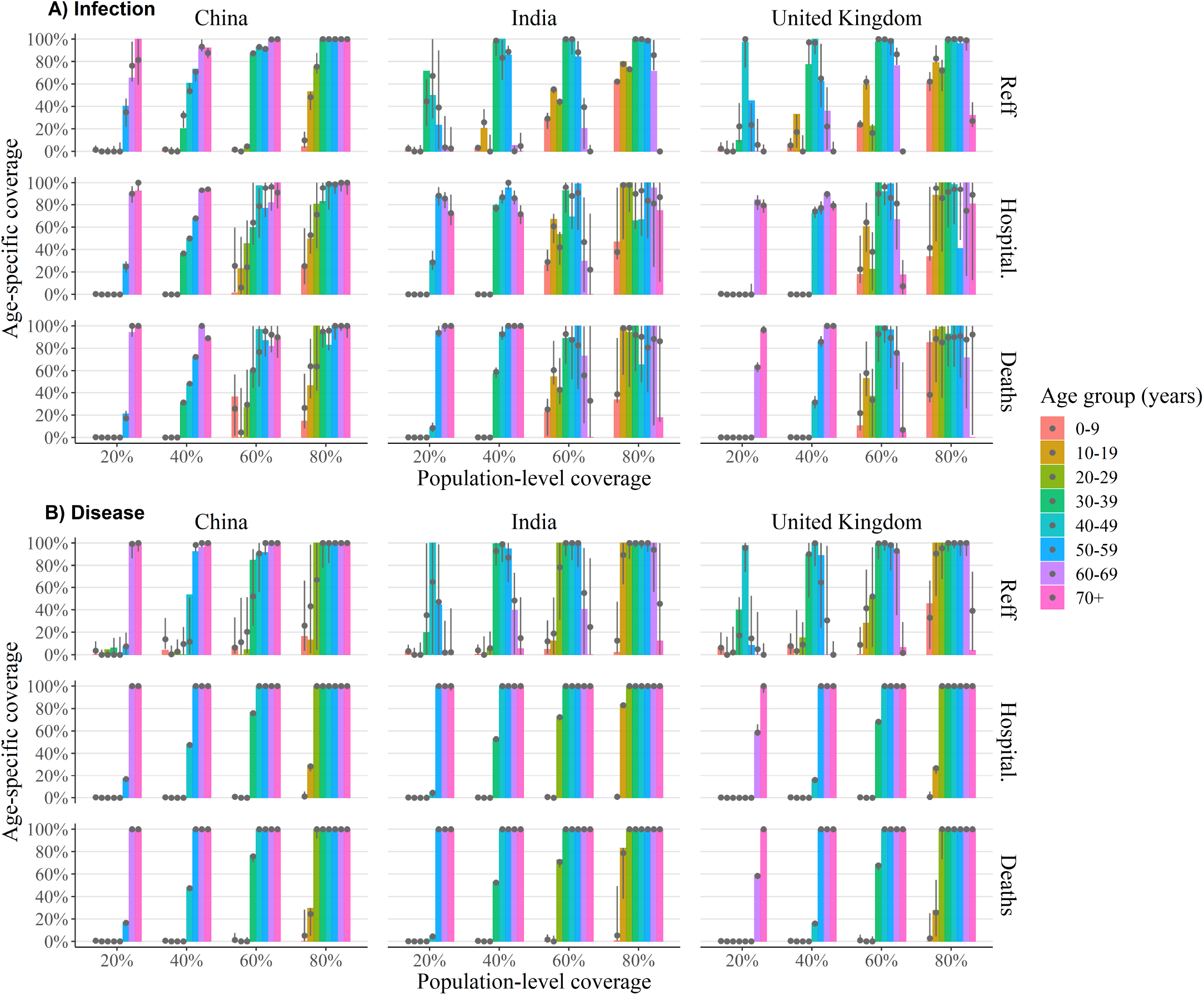
Optimal age-specific vaccination policy, allowing age-dependent clinical fraction only (M3). The optimum coverage level for each age group (coloured bars) for infection-preventing (**A**) and disease-preventing (**B**) vaccines for varying population coverage levels to minimize transmission (top), hospitalizations (middle) and deaths (bottom) for India (left), China (middle) and the United Kingdom (right). Alongside the global optimal solution (coloured bars) we provide the 2.5 − 97.5 percentile ranges (vertical grey lines) and median values (grey dots) of age-specific coverage levels for all locally optimal solutions whose target value is within 1% of the global optimal value. Here we have assumed that the vaccine is 90% effective across all age groups (remaining model parameters assume their baseline values which can be found in Table S1).

## Notes

### Competing Interest Statement

The authors have declared no competing interest.

### Summary of Updates

Analysis updated to incorporate: hospitalizations and deaths as additional optimization targets; updated estimates of the vaccine efficacy; and further sensitivity analysis on the relative transmissibility of asymptomatic transmitters.

